# A rare splice-site variant in cardiac troponin-T (*TNNT2)*: The need for ancestral diversity in genomic reference datasets

**DOI:** 10.1101/2024.02.08.24302375

**Authors:** Alexandra Butters, Kate Thomson, Franki Harrington, Natasha Henden, Karen McGuire, Alicia B. Byrne, Samantha Bryen, Kathryn A. McGurk, Megan Leask, Michael J. Ackerman, John Atherton, Johan M. Bos, Colleen Caleshu, Sharlene Day, Kyla Dunn, Ian Hayes, Jimmy Juang, Julie McGaughran, Natalie Nowak, Victoria N. Parikh, Anne Ronan, Christopher Semsarian, Jil C. Tardiff, Marianne Tiemensma, Tony R. Merriman, James S. Ware, Jonathan R. Skinner, Daniel G. MacArthur, Owen M. Siggs, Richard D. Bagnall, Jodie Ingles

**Author notes:** **ADDRESS FOR CORRESPONDENCE,** Associate Professor Jodie Ingles, Genomics and Inherited Disease Program, Garvan Institute of Medical Research, 384 Victoria Street Darlinghurst, Sydney, New South Wales 2010 Australia Twitter: @jodieingles.

## Abstract

The underrepresentation of different ancestry groups in large genomic datasets creates difficulties in interpreting the pathogenicity of monogenic variants. Genetic testing for individuals with non-European ancestry results in higher rates of uncertain variants and a greater risk of misclassification. We report a rare variant in the cardiac troponin T gene, *TNNT2*; NM_001001430.3: c.571-1G>A (rs483352835) identified via research-based whole exome sequencing in two unrelated probands of Oceanian ancestry with cardiac phenotypes.

The variant disrupts the canonical splice acceptor site, activating a cryptic acceptor and resulting in an in-frame deletion (p.Gln191del). The variant is rare in gnomAD v4.0.0 (13/780,762; 0.002%), with the highest frequency in South Asians (5/74,486; 0.007%) and has 16 ClinVar assertions (13 diagnostic clinical laboratories classify as variant of uncertain significance). There are at least 28 reported cases, many with Oceanian ancestry and diverse cardiac phenotypes. Indeed, among Oceanian-ancestry-matched datasets, the allele frequency ranges from 2.9-8.8% and is present in 2/4 (50%) Indigenous Australian alleles in Genome Asia 100K, with one participant being homozygous. With Oceanians deriving greater than 3% of their DNA from archaic genomes, we found c.571-1G>A in Vindija and Altai Neanderthal, but not the Altai Denisovan, suggesting an origin post Neanderthal divergence from modern humans 130-145 thousand years ago. Based on these data, we classify this variant as benign, and conclude it is not a monogenic cause of disease. Even with ongoing efforts to increase representation in genomics, we highlight the need for caution in assuming rarity of genetic variants in largely European datasets. Efforts to enhance diversity in genomic databases remain crucial.

## INTRODUCTION

Inherited cardiomyopathies such as hypertrophic cardiomyopathy (HCM), dilated cardiomyopathy (DCM) and restrictive cardiomyopathy (RCM) cumulatively affect ∼1 in 200-500 in the general population.^1^ These conditions often have an autosomal dominant inheritance pattern with marked clinical and genetic heterogeneity. Some patients report only mild symptoms, while others experience more severe outcomes such as heart failure or sudden cardiac death (SCD). Genetic testing of genes with established clinical validity can identify the causal variant in approximately half of these patients and is part of mainstream clinical management.^2^ Genetic variations encoding the cardiac sarcomere proteins, such as *TNNT2* (troponin T), are an important cause.^2,3^ *TNNT2* encodes cardiac troponin T, an integral protein in the cardiac sarcomere. The troponin complex and calcium regulate the binding of actin and myosin. *TNNT2* is definitively associated with HCM and DCM.^4,5^ While causative variants in *TNNT2* have been reported in cases of RCM, the RCM gene-disease association has not been formally curated.

The underrepresentation of individuals from diverse ancestry groups in population genomic datasets creates challenges in interpreting potentially causal monogenic variants.^6^ As a result, genetic testing has a lower diagnostic yield for people not of European ancestry due to more variants being considered of uncertain significance.^7,8^ In the context of rare monogenic diseases, population frequency is important when considering whether a variant is causal, with disease prevalence, penetrance, and known genetic heterogeneity informing an acceptable allele frequency for pathogenicity in a specific disease.^9,10^ However, while a genetic variant may appear to be rare in large population reference databases such as the Genome Aggregation Database (gnomAD),^11^ if the person being tested has a genetic ancestry that is not well represented, then there should be caution in considering whether the rarity criterion is met.^12^

There is a concerted international effort to increase ancestral diversity in genomics, with gnomAD v4.0.0 now including an additional 138,000 individuals from previously underrepresented backgrounds and refined recognition of genetic ancestry groups.^13^ However, individuals with genetic ancestry derived from Oceania remain virtually absent from genomic databases. Oceania is a geographical region on the Pacific Ocean comprising Australasia, Melanesia, Micronesia and Polynesia.^13^ The history of modern humans in Oceania comprises two vastly divergent ancestral populations. The first arrived in Southeast Asia more than 40,000 years ago, contributing to the ancestry of Papuans, Indigenous Australians and some Pacific Island populations.^14–16^ The second closely relates to mainland East Asians, specifically Indigenous Taiwanese, who populated remote Oceania ∼3,000 years ago.^16,17^

We report a variant, *TNNT2*; NM_001001430.3: c.571-1G>A (rs483352835), in two unrelated probands with cardiac phenotypes and Oceanian ancestry. Given its relative rarity in available population databases but the prevalence of Oceanian ancestry in reported cases, we aimed to evaluate it as a cause of monogenic cardiomyopathy.

## METHODS

*TNNT2*; NM_001001430.3: c.571-1G>A (rs483352835) was identified in two unrelated probands seen at a specialised genetic heart disease clinic, Sydney, Australia, who consented to research-based whole genome sequencing (Royal Prince Alfred Hospital Sydney Local Health District Human Research Ethics Committee X15-0089). The variant classification was performed by research genetic counsellors using the *MYH7-*modified ACMG/AMP criteria and discussed at a dedicated cardiac genetic multidisciplinary team meeting.^9^

To identify probands with this variant, literature and ClinVar reports for the c.571-1G>A (rs483352835) variant were sought, and clinical laboratories and research groups were contacted for additional information about patients’ cardiac phenotypes and ancestry.

The frequency of the variant in open-access genomic databases, including gnomAD v3.12 and v4.0.0, Genome Asia 100K (Oceania), and the Regeneron Genetics Center Mexico City Prospective Study Browser, n=140,000, was accessed. We then contacted research groups with ancestry-matched participants with non-cardiac diseases and available genomic data. This included the Taiwan Biobank (n=1517), which consists primarily of Han Chinese and does not include any Indigenous Taiwanese individuals;^18^ the Pacific Islands Rheumatic Heart Disease Genetics Network (n=3234) which consists of low-coverage genomes and SNP arrays with imputation^19^ and includes participants from Melanesia (Vanuatu, New Caledonia, Fiji), West Polynesia (Tonga, Samoa), and East Polynesia (Cook Islands, French Polynesia); and a group of 194 people of self-reported Aotearoa New Zealand Māori or Pacific Islander ethnicity who had previously been recruited and consented according to the Health and Disability Ethics approval (MEC/05/10/130) were genotyped to determine the allele frequency of the *TNNT2* c.571-1G>A (rs483352835) variant in these populations. Genotyping was carried out by sequencing as described in Emde et al. 2021.^20^

## RESULTS

### Patient summary

Proband 1 was a male of Oceanian ancestry who suffered an SCD in his early 20s with no cause identified following a comprehensive post-mortem investigation. Proband 2 was a 0-3 year-old child of Oceanian ancestry with RCM who died from rapidly progressive heart failure; in addition to the *TNNT2* splice variant, a pathogenic variant in *TNNI3* classified pathogenic as per ACMG guidelines was also identified.

We queried ClinVar and the literature to establish if the variant had been reported previously. There were 16 ClinVar entries for this variant (ClinVar variation ID: 132940), updated between December 2014 to January 2024. Fourteen entries classify this variant as being of uncertain significance due to haploinsufficiency not being established as a disease mechanism for *TNNT2*. One proposed this variant as likely pathogenic in 2010, primarily due to the absence of *TNNT2* splice variants among available controls. The final entry gives no classification. Of the 16 submitters, 13 are diagnostic genetic laboratories.

### Case data

Case data were collated from ClinVar entries, biomedical literature, and specialized cardiovascular genetic disease centers, giving 26 unrelated probands with this variant (excluding our two probands). Of these, 24 had a known cardiac phenotype, including 15 with HCM (including two diagnosed on autopsy following SCD), five with DCM (two presented at less than three years of age),^5^ three with sudden unexplained death, and one individual with high-burden premature ventricular contractions, syncope, and a family history of SCD. One individual lacked a specific diagnosis but had a family history of SCD, and in another, the diagnosis was not available. Four probands reported a family history of SCD in a close relative. Evidence of co-segregation with disease in families was reported for one family with HCM, with the variant identified in two affected relatives. At least four probands (15%) had an additional pathogenic variant that explained their cardiac phenotype. Ancestry was available for 22 probands, of which 21 probands were reported as having Oceanian ancestry, including Aotearoa New Zealand Māori, Samoan, Tongan, Polynesian, Pacific Islander, Australian Aboriginal and Torres Strait Islander, and Hawaiian (Figure 2).

### Variant summary

*TNNT2* exon numbering is in reference to the NM_001001430.3 transcript. The c.571-1G>A (rs483352835) variant disrupts the canonical acceptor splice site of exon 12, an alternatively spliced exon that contains just three amino acids and is expressed in 7/13 of NCBI RefSeq transcripts^21^ (Figure 1A). SpliceAI^22^ predicts the c.571-1G>A (rs483352835) variant abolishes the canonical acceptor and strengthens a cryptic acceptor three nucleotides downstream (Figure 1B). RNA studies have shown that the variant results in activation of the cryptic acceptor, leading to an in-frame deletion of one amino acid, p.Gln191del.^4^ The activated cryptic acceptor is an annotated acceptor splice site in NM_001001432.3 (Figure 1C), and use of this acceptor has been observed in 25% of RNA sequencing samples in SpliceVault, a dataset comprised of Genotype-Tissue Expression (GTEx) data and Sequence Read Archive (SRA) data.^23^ Proportion expressed across transcripts (pext) scores^24^ suggest that exon 12 expression is reduced relative to other exons in *TNNT2*, supporting alternative splicing of this exon. The three amino acids encoded by this exon are weakly conserved, with no pathogenic/likely pathogenic missense variants reported in ClinVar. These data suggest that the p.Gln191del is a common, tolerated event. The c.571-1G>A (rs483352835) variant is likely to increase the proportion of transcripts with p.Gln191del, an event likely to have minimal impact on *TNNT2* expression and/or function.

**Figure 1:**
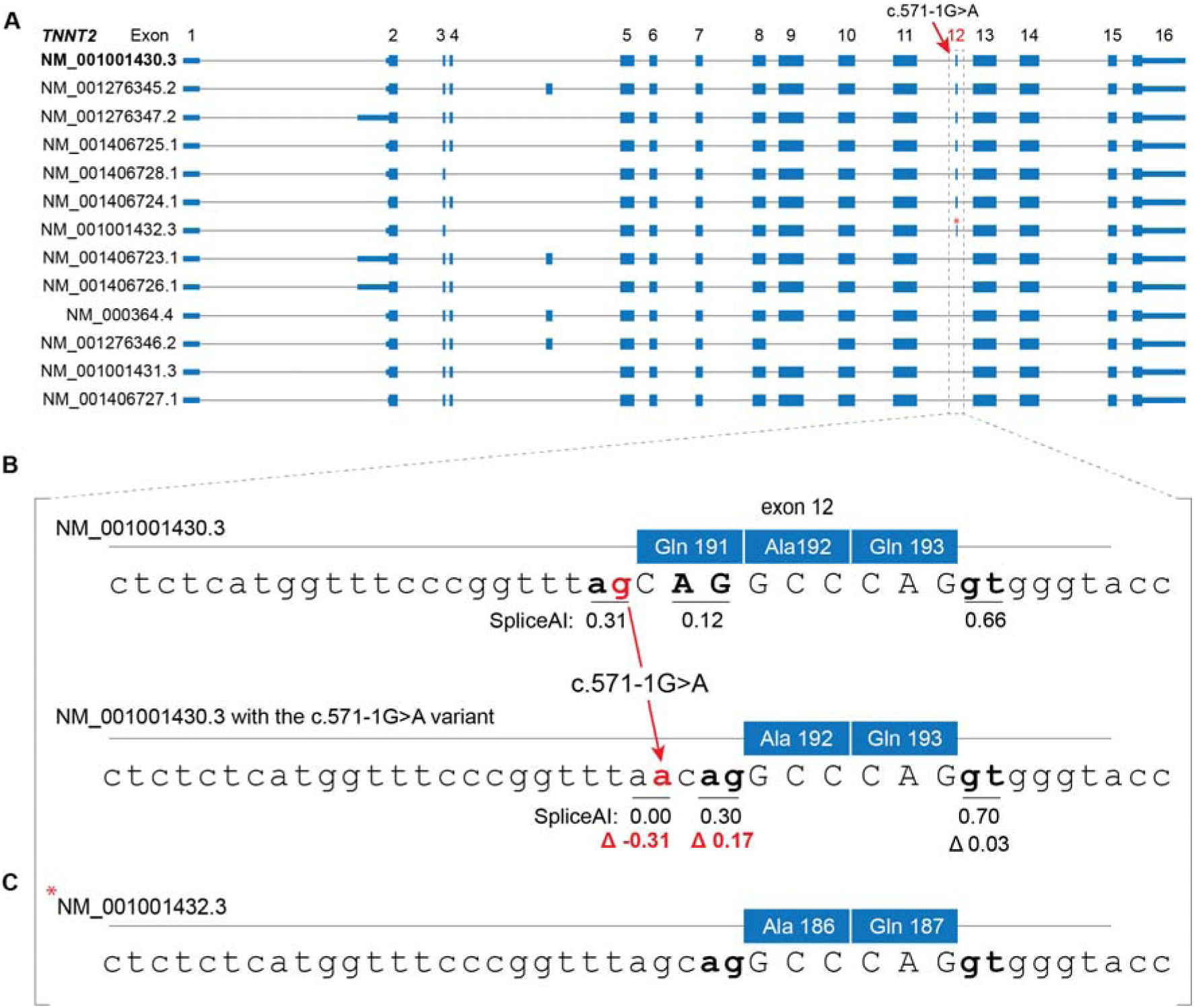
**A:** NCBI RefSeq transcripts for the *TNNT2* gene. Exons are numbered as per NM_001001430.3. **B:** SpliceAI predicts weakening of the canonical acceptor splice site in exon 12 and strengthening of a cryptic acceptor splice site 3 nucleotides downstream, resulting in deletion of Gln191. There is no significant change in SpliceAI score for the exon 12 canonical donor splice site. **C:** The cryptic acceptor is the annotated acceptor splice site for NM_001001432.3.

**Figure 2:**
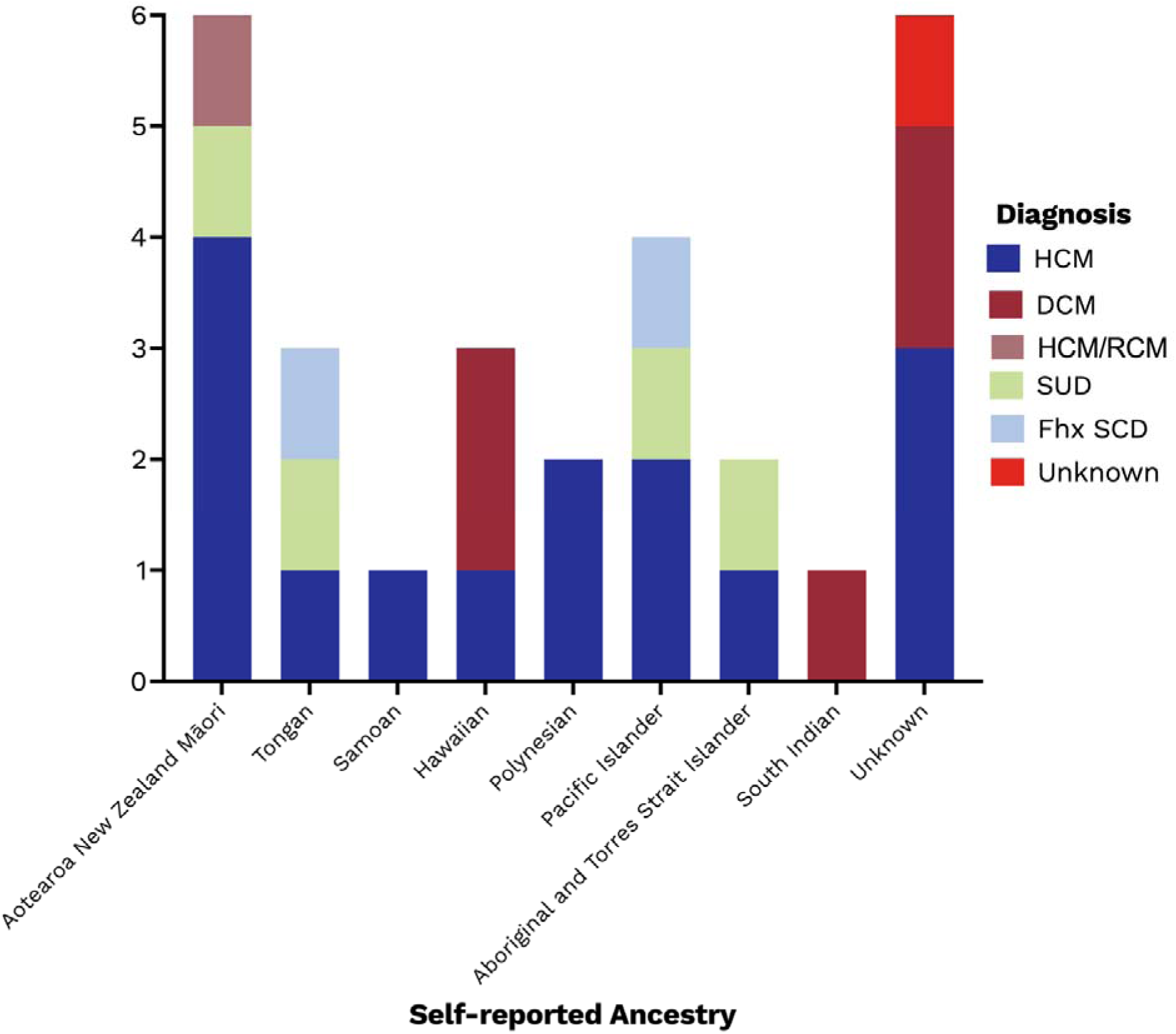
Probands with the *TNNT2* c.571-1G>A (rs483352835) splice-site variant by self-reported ancestry and disease phenotype. Abbreviations: HCM, hypertrophic cardiomyopathy; DCM, dilated cardiomyopathy; RCM, restrictive cardiomyopathy; SUD, sudden unexplained death; Fhx, family history; SCD, sudden cardiac death.

### Frequency in population databases

Allele frequencies are shown in Table 1. Among publicly available databases, *TNNT2*:c.571-1G>A (rs483352835) was present in gnomAD v4.0.0 in 13/780,762 (0.002%) alleles, with the highest sub-population frequencies among South Asian (5/74,486; 0.007%) and East Asian (1/41,260; 0.002%) alleles.^11^ Within Genome Asia 100K Project, the variant was present in 6/148 (4.1%) chromosomes in the Oceanian population, with a frequency of 2/4 (50%) chromosomes in the 2 Australian participants (one homozygote), and 4/140 (2.9%) in Papuan alleles (Figure 3).^25^ The variant was not present in the Regeneron Genetics Center Mexico City Prospective Study Browser (0/301,992; 0%).^26^

**Figure 3:**
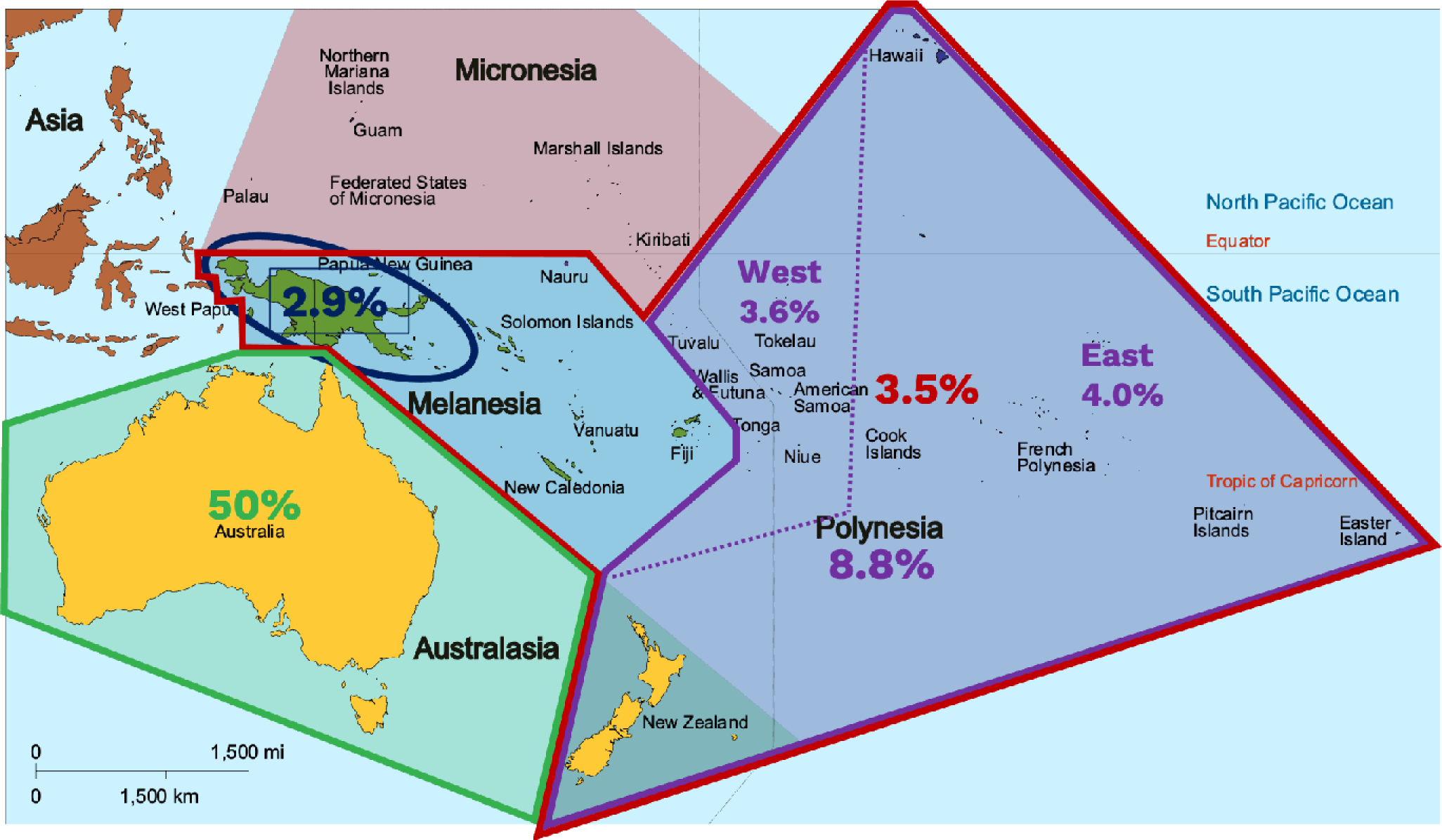
Geographical Map of Oceania (Polynesia, Micronesia and Melanesia) with population-specific *TNNT2* c.571-1G>A (rs483352835) allele frequencies. Note the 50% allele frequency in Indigenous Australians is based on 2 individuals. This work is licensed under Attribution-ShareAlike 3.0 Unported (CC BY-SA 3.0). The original can be found here: https://commons.wikimedia.org/wiki/File:Oceania_UN_Geoscheme_-_Map_with_Zones.svg#/media/File:Oceania_UN_Geoscheme_Regions.

**TABLE 1:** Frequency of Variant *TNNT2*; NM_001001430.2: c.571-1G>A (rs483352835) in genomic reference databases.

| Reference Group | Data availability | Allele count | Allele Frequency | % |
| --- | --- | --- | --- | --- |
| **gnomAD v3.1.2** | Open | 2/152,188 | 0.000013 | 0.001 |
| **gnomAD v4.0.0** | Open | 13/780,762 | 0.000017 | 0.002 |
| Genetic ancestry group: Remaining individuals |  | 6/35,206 | 0.00017 | 0.017 |
| Genetic ancestry group: South Asian |  | 5/74,486 | 0.00007 | 0.007 |
| Genetic ancestry group: East Asian |  | 1/41,260 | 0.00002 | 0.002 |
| **Genome Asia 100K (Oceania)** | Open | 6/148 | 0.041 | 4.1 |
| Australia (includes 1 homozygote) |  | 2/4 | 0.5 | 50 |
| Papua New Guinea |  | 4/140 | 0.029 | 2.9 |
| **Regeneron Genetics Center Mexico City Prospective Study Browser, n=150,996** | Open | 0/301,992 | 0 | 0 |
| **Taiwan Biobank (primarily Han Chinese)** | On request | 0/3032 | 0.0 | 0.0 |
| **Pacific Islands Rheumatic Heart Disease Genetics Network, n=3,234** | On request |  |  |  |
| Polynesian subgroup |  | - | 0.088 | 8.8 |
| Broader (including Melanesian) |  | - | 0.035 | 3.5 |
| **Māori and Pacific Islander populations of Aotearoa New Zealand, n=194 controls** | On request |  |  |  |
| East Polynesian subgroup (n=139) |  | - | 0.039 | 4.0 |
| West Polynesian subgroup (n=55) |  | - | 0.036 | 3.6 |

We determined the presence of the variant within large biomedical research studies, including All of Us and the UK Biobank. In All of Us (n=245,388), the variant was found in fewer than 20 individuals, none of whom reported a diagnosis of cardiomyopathy, noting that occurrences <20 must be reported as such. Among these carriers, 59% were identified as Native Hawaiian or Other Pacific Islander.^27^ In the UK Biobank, out of 469,835 individuals who underwent exome sequencing, only one participant, who identified as belonging to the “Other ethnic group” and was born in South-East Asia, had the variant. Notably, this participant did not have cardiomyopathy.

Among ancestry-relevant population sequencing data unavailable in the public domain, the variant was absent in the Taiwanese Biobank (0/3032; 0%). Within the Pacific Islands Rheumatic Heart Disease Genetics Network, the variant was present at a frequency of 0.088 (8.8%) within the Polynesian sub-group and 0.035 (3.5%) across the wider group, including Melanesians and South Asians. In a genome sequencing dataset (low-pass sequencing followed by imputation),^20^ of participants of Aotearoa New Zealand Māori and Pacific Islander ancestry recruited to a Health and Disability study, the variant was present at an allele frequency of 4.0% in East Polynesian individuals (n=139), and 3.6% among West Polynesian individuals (n=55) (Figure 3).

### Presence in archaic genomes

Of note, indigenous Papuan and Australian people derive greater than 3% of their DNA from Neanderthal ancestry,^28^ which is a higher percentage than for Eurasian populations.^28,29^ *TNNT2*:c.571-1G>A (rs483352835) was shown to be present in two archaic genomes (Vindija and Altai Neanderthal), but not the Altai Denisovan,^28–31^suggesting it might have arisen ≥130-145 thousand years ago after the Neanderthal populations diverged from modern humans.^20^ This likely explains the increased frequency of *TNNT2*:c.571-1G>A (rs483352835) in Oceanian populations.

### Benign variant classification

The ACMG/AMP guidelines state that a minor allele frequency of greater than 0.05 for an autosomal dominant gene can be considered stand-alone evidence of benign impact (BA1 criterion).^32^ Taking into consideration the allele frequencies among individuals with Oceanian ancestry, we applied the BA1 criterion and classify *TNNT2*:c.571-1G>A (rs483352835) as a benign variant, which is predicted to lead to a tolerated in-frame deletion of a single amino acid.

## DISCUSSION

The limited representation of individuals with non-European ancestry in genetic studies and genomics reference datasets poses challenges in accurately classifying and interpreting genetic variants identified in underrepresented and Indigenous populations. This disparity results in a 2-3 times higher risk of uncertain variant identification following genetic testing for non-European individuals,^33,34^ compromising the utility of such testing and increasing the likelihood of variant misclassification.^35^ We demonstrate the importance of publicly accessible ancestry-matched populations in enabling accurate variant interpretation, thereby avoiding potential harms that would disproportionately impact individuals from historically underrepresented ancestries. In this example, the *TNNT2*:c.571-1G>A (rs483352835) genetic variant has been frequently identified by testing laboratories and in combination with numerous case reports, could be misinterpreted as causative of disease. Indeed, many diagnostic laboratory summaries in ClinVar note the lack of evidence of *TNNT2* haploinsufficiency as a disease mechanism as the barrier to considering this variant as causative.

Contemporary variant classification evaluates all reported variant cases under assessment and considers the individuals’ clinical phenotype. Our experience highlights the need for patient ancestry to be a key data field in these assessments. Due to the scarcity of ancestry-matched reference data for Oceanians, our only course of action was to identify and personally contact laboratories and research groups with sequencing data from Oceanian peoples, ultimately revealing that the variant is too common to be considered disease-causing. As a result, despite some evidence that might support pathogenicity, the ancestry-matched allele frequency indicates this to be a benign variant. Given that 14 ClinVar assertions from clinical diagnostic laboratories report this as a VUS, this classification downgrade will ensure it is not reported and any future time or resources aimed at reclassification are carefully considered. While we cannot exclude a potential modifier role in disease, this variant is not a highly penetrant monogenic cause of cardiomyopathy but is a common single nucleotide variant within Oceanian populations.

Efforts to increase ancestral diversity in genomics have produced larger and more inclusive publicly available reference databases, yet there is a considerable way to go in achieving global representation.^13,20,36^ Efforts to improve representation in genomic databases rely heavily on effective community engagement, and partnerships with underrepresented groups are essential.^37^ Open sharing of these data in ways that comply with participant consent, ethical standards and community governance will enable a more accurate understanding of the genetic basis of health and disease.^38^ While there are important considerations when requesting patients to disclose their ancestry, in the setting of genetic testing and variant interpretation, these data are critical for avoiding misclassification. Moreover, in cases where this information is unavailable, inferring ancestry from genetic tests becomes essential, as it can effectively prevent the generation of undue anxiety in patients and help mitigate the risk of unnecessary medical interventions.^39^

## CONCLUSION

We highlight the challenges of interpreting ‘rare’ monogenic variants among individuals from poorly represented ancestry groups. Our analysis of *TNNT2:*c.571-1G>A (rs483352835), a common single nucleotide variant within Oceanian populations, emphasizes the critical need for openly accessible, large and diverse genomic reference databases to ensure accurate interpretation of variants and the value of genetic testing. While achieving ancestral diversity will require significant time, commitment, and resources, including deep community engagement and addressing existing barriers to research participation and mistrust,^40^ it is a necessary step if we are to ensure the entire population can benefit equitably from genomic medicine.

## DATA AVAILABILITY

Data supporting this paper are contained within the article. Any additional data will be available upon reasonable request and following appropriate ethical and governance approvals.

## ETHICS STATEMENT

This study adheres to the principles set out in the Declaration of Helsinki.

Ethics approval was obtained in accordance with policies applicable at Royal Prince Alfred Hospital Sydney Local Health District Human Research Ethics Committee (X15-0089 and X17-0350) for the original two patients, who provided written informed consent, and for collection of de-identified summary data from clinical laboratories and research centers used to inform variant prioritization and classification. Ethical approval for other summary case data including written informed consent from participants was granted by Melbourne Health Human Research Ethics Committee and New Zealand Ethics Committee. Ethical approval with a waiver of consent was granted by Stanford School of Medicine, USA; Pennsylvania University Medical Center, USA. All other data were in the public domain. The UK Biobank analysis (National Research Ethics Service, 11/NW/0382) was conducted under the terms of access of project 47602. The All of US analysis (v7) was conducted through workspace 9c419818.

## REFERENCES

1. Semsarian, C., Ingles, J., Maron, M. S. & Maron, B. J. New perspectives on the prevalence of hypertrophic cardiomyopathy. J Am Coll Cardiol 65, 1249–54 (2015).

2. Ingles, J., et al. Evaluating the Clinical Validity of Hypertrophic Cardiomyopathy Genes. Circ Genom Precis Med 12, e002460 (2019).

3. Jordan, E. et al. Evidence-Based Assessment of Genes in Dilated Cardiomyopathy. Circulation 144, 7–19 (2021).

4. Van Driest, S. L. et al. Comprehensive Analysis of the Beta-Myosin Heavy Chain Gene in 389 Unrelated Patients With Hypertrophic Cardiomyopathy. J. Am. Coll. Cardiol. 44, 602–610 (2004).

5. Rani, D. S., Dhandapany, P. S., Nallari, P., Narasimhan, C. & Thangaraj, K. A novel arginine to tryptophan (R144W) mutation in troponin T (cTnT) gene in an indian multigenerational family with dilated cardiomyopathy (FDCM). PloS One 9, e101451 (2014).

6. Sirugo, G., Williams, S. M. & Tishkoff, S. A. The Missing Diversity in Human Genetic Studies. Cell 177, 26–31 (2019).

7. Petrovski, S. & Goldstein, D. B. Unequal representation of genetic variation across ancestry groups creates healthcare inequality in the application of precision medicine. Genome Biol. 17, 157 (2016).

8. Bustamante, C. D., De La Vega, F. M. & Burchard, E. G. Genomics for the world. Nature 475, 163–165 (2011).

9. Richards, S. et al. Standards and Guidelines for the Interpretation of Sequence Variants: A Joint Consensus Recommendation of the American College of Medical Genetics and Genomics and the Association for Molecular Pathology. Genet. Med. Off. J. Am. Coll. Med. Genet. 17, 405–424 (2015).

10. Whiffin, N. et al. Using high-resolution variant frequencies to empower clinical genome interpretation. Genet. Med. 19, 1151–1158 (2017).

11. Karczewski, K. J. et al. The mutational constraint spectrum quantified from variation in 141,456 humans. Nature 581, 434–443 (2020).

12. ClinGen Sequence Variant Interpretation Recommendation for PM2 - Version 1.0. (2020).

13. Koenig, Z. et al. A harmonized public resource of deeply sequenced diverse human genomes. BioRxiv Prepr. Serv. Biol. 2023.01.23.525248 (2023) doi:10.1101/2023.01.23.525248.

14. Malaspinas, A.-S. et al. A genomic history of Aboriginal Australia. Nature 538, 207–214 (2016).

15. Rasmussen, M. et al. An Aboriginal Australian genome reveals separate human dispersals into Asia. Science 334, 94–98 (2011).

16. Wollstein, A. et al. Demographic History of Oceania Inferred from Genome-wide Data. Curr. Biol. 20, 1983–1992 (2010).

17. Skoglund, P. et al. Genomic insights into the peopling of the Southwest Pacific. Nature 538, 510– 513 (2016).

18. Feng, Y.-C. A. et al. Taiwan Biobank: A rich biomedical research database of the Taiwanese population. Cell Genomics 2, 100197 (2022).

19. Parks, T. et al. Association between a common immunoglobulin heavy chain allele and rheumatic heart disease risk in Oceania. Nat. Commun. 8, 14946 (2017).

20. Emde, A.-K. et al. Mid-pass whole genome sequencing enables biomedical genetic studies of diverse populations. BMC Genomics 22, 666 (2021).

21. O’Leary, N. A. et al. Reference sequence (RefSeq) database at NCBI: current status, taxonomic expansion, and functional annotation. Nucleic Acids Res. 44, D733–745 (2016).

22. Jaganathan, K. et al. Predicting Splicing from Primary Sequence with Deep Learning. Cell 176, 535–548.e24 (2019).

23. Dawes, R. et al. SpliceVault predicts the precise nature of variant-associated mis-splicing. Nat. Genet. 55, 324–332 (2023).

24. Cummings, B. B. et al. Transcript expression-aware annotation improves rare variant interpretation. Nature 581, 452–458 (2020).

25. Wall, J. D. et al. The GenomeAsia 100K Project enables genetic discoveries across Asia. Nature 576, 106–111 (2019).

26. Ziyatdinov, A. et al. Genotyping, sequencing and analysis of 140,000 adults from Mexico City. Nature 622, 784–793 (2023).

27. Bick, A. G. et al. Genomic data in the All of Us Research Program. Nature 627, 340–346 (2024).

28. Prüfer, K. et al. A high-coverage Neandertal genome from Vindija Cave in Croatia. Science 358, 655–658 (2017).

29. Yuan, K. et al. Refining models of archaic admixture in Eurasia with ArchaicSeeker 2.0. Nat. Commun. 12, 6232 (2021).

30. Prüfer, K. et al. The complete genome sequence of a Neanderthal from the Altai Mountains. Nature 505, 43–49 (2014).

31. Reich, D. et al. Denisova Admixture and the First Modern Human Dispersals into Southeast Asia and Oceania. Am. J. Hum. Genet. 89, 516–528 (2011).

32. Ghosh, R. et al. Updated recommendation for the benign stand-alone ACMG/AMP criterion. Hum. Mutat. 39, 1525–1530 (2018).

33. Butters, A. et al. Clinical Profile and Health Disparities in a Multiethnic Cohort of Patients With Hypertrophic Cardiomyopathy. Circ. Heart Fail. 14, e007537 (2021).

34. Stafford, F. et al. The role of genetic testing in diagnosis and care of inherited cardiac conditions in a specialised multidisciplinary clinic. Genome Med 14, 145 (2022).

35. Manrai, A. K. et al. Genetic Misdiagnoses and the Potential for Health Disparities. N Engl J Med 375, 655–65 (2016).

36. Mapes, B. M. et al. Diversity and inclusion for the All of Us research program: A scoping review. PloS One 15, e0234962 (2020).

37. Lemke, A. A. et al. Addressing underrepresentation in genomics research through community engagement. Am. J. Hum. Genet. 109, 1563–1571 (2022).

38. Hudson, M. et al. Rights, interests and expectations: Indigenous perspectives on unrestricted access to genomic data. Nat. Rev. Genet. 21, 377–384 (2020).

39. Appelbaum, P. S. et al. Is there a way to reduce the inequity in variant interpretation on the basis of ancestry? Am. J. Hum. Genet. 109, 981–988 (2022).

40. Sharma, A. & Palaniappan, L. Improving diversity in medical research. Nat. Rev. Dis. Primer 7, 74 (2021).

